# Is health expenditure on immunisation associated with immunisation coverage in Sub-Saharan African countries?

**DOI:** 10.1101/2022.07.31.22278245

**Authors:** Israel Oluwaseyidayo Idris, Janet Tapkigen, Luke Ouma, Francis Ifeanyi Ayomoh, Gabriel Omoniyi Ayeni

## Abstract

In a bid to address the high burden of vaccine-preventable disease and low immunisation coverage in Africa, Ministers of health and finance from several African countries conveyed at the maiden Ministerial Conference on Immunisation at Addis Ababa, Ethiopia on the 25th of February 2016 to pledge political commitments to reduce the prevalence and deaths from vaccine-preventable diseases to the barest minimum. The fulfilment of this pledge across Africa would require the design of contextually tailored sustainable plans to finance the procurement of vaccines and the running of apt immunisation programs. A robust understanding of the trend of immunisation financing in Africa will support the development of suitable national immunization financing plans, guide policy makers to develop immunisation financing strategies focused on domestic resources but factor in donor support; and provide insights for the rejuvenation and expansion of immunisation programs. Our study’s objective is to estimate the minimum fraction of a country’s health budget that should be invested in the national immunisation programme to achieve a national immunisation coverage of 80% or greater depending on the context with and without donors’ support.

The study results did not find any evidence to indicate that health expenditure on immunisation (as a proportion of total health expenditure) could be a strong predictor of immunisation coverage. However, we observed an association between total health expenditure (as a % of the GDP) and immunisation coverage, for BCG (p=0.047) and DPT3 (p=0.013) vaccines. Therefore, health expenditure as a percentage of GDP can be considered as an important predictor of immunisation coverage. We demonstrate in selected countries that to achieve the GAVI target of 80% in the countries with low DPT3 coverage, health expenditure as a percentage of GDP would need to be increased by more than 45%. We are optimistic that our study results and recommendations will facilitate the development of strategies that support African countries to increase domestic financing for national immunization programmes towards achieving 2030 targets for immunization coverage.

**Evidence before this study:** We conducted a desk review to identify official government records and reports on immunisation financing in African countries in Google scholar, WHO Library, GAVI and World bank databases using keywords such as “immunisation financing”, “health budget”, “health financing policies”, “immunisation financing policies” and “health expenditure”. We identified data for all countries in Africa but were only able to retrieve complete data from 24 countries. We considered the retrieved data for each country to be complete for our study if we found data on immunisation expenditure, health expenditure as a percentage of Gross Domestic Product, Gross Domestic Product, BCG coverage, DPT3 coverage, PCV1 coverage, MCV1 coverage, fertility rates, under-five mortality rates, under-five population and the total population.

**Added value of this study:** We sought for any association between immunisation expenditure and health expenditure (as a % of the GDP) and immunisation coverage over a five-year period (2013 to 2017) in twenty-four African countries. To our knowledge, this is the first study that has shown a correlation between immunisation financing, health expenditure and immunisation coverage and how this association varies across countries. Prediction modelling of vaccine coverage time series for countries with less than desired level of coverage (below 80%) enabled us to construct a predictive index that visualised the effect of increasing health expenditure (as a % of the GDP) would have on immunisation coverage with all other variables unchanged.

**Implications of the available evidence:** We posit that immunisation expenditure is not a statistically significant predictor for immunisation coverage for DPT3 and BCG vaccines; rather, with strong statistical evidence, health expenditure (as a % of the GDP) can be used to predict immunization coverage. Our prediction model estimated the percentage increase in health expenditure (as a % of the GDP) that would be required for countries with low immunization coverage to attain the target for immunization coverage recommended by the IA2030 Framework for Action.

## Introduction

“No child should die or be sickened by vaccine-preventable diseases’’, a phrase coined and pledged during the 1st Ministerial Conference on Immunization in Africa, held from 24-25 February 2016 in Addis Ababa, Ethiopia (1). At the time of the 2016 declaration on immunization, close to a million African children – inclusive of new-borns, die before their fifth birthday from vaccine preventable diseases annually; and every year, about 30 million children under five years of age get sick from vaccine-preventable diseases (VPDs) in Africa (2,3). While some of the commitments to achieve the above pledge in the conference included strengthening supply chains and delivery systems, increasing universal access to vaccines and sustainable immunisation financing, the latter remains the most critical and can be arduous to achieve (1). This is because international donors are gradually reducing their funding to immunisation programs in low- and middle-income countries (LMICs); as other health priorities, such as Human immunodeficiency virus infection and acquired immunodeficiency syndrome (HIV/AIDS) programmes, compete for limited health funding (4). Additionally, the financial resources needed for the successful implementation of immunisation programmes in LMICs are increasing because of high costs associated with the provision of the recent World Health Organisation (WHO) recommended vaccines such as the pneumococcal conjugate vaccine (PCV) and the cost of extending immunisation activities to hard-to-reach areas (5). As Africa countries struggle to finance their national immunization programmes which require more resources amid dwindling donor funding, understanding the trend of immunisation financing in these countries would assist in the development of appropriately tailored country-level financing strategies that would advance sustainable domestic financing for immunization programmes and support the expansion of these programmes to optimize national immunization coverage.

This study is aimed at critically analysing immunisation financing in Sub-Saharan African countries from 2013-2017. With strong statistical evidence, our study’s objective is to estimate the minimum percentage of a country’s health budget - with and without donors’ support - that must be invested in the national immunisation programmes with the aim to achieve a national immunisation coverage of 80% - 90% depending on the contexts

## Methodology

### Data Description

Sub-Saharan Africa consists of 23 low-income, 18 lower-middle income and 4 upper-middle income countries according to the 2022 World Bank classification (6). This paper focuses on 17 low-income countries: Burkina Faso, Burundi, Ethiopia, Gambia, Ghana, Madagascar, Mali, Mozambique, Rwanda, Sierra Leone, Sudan, Togo, Central Africa Republic, Chad, Republic of the Congo, Eritrea, Guinea, and Liberia and 7 lower-middle income countries: Benin, Côte D’Ivoire, Djibouti, Senegal, Zambia, and Zimbabwe. Of the 24 countries in this study, all are in the Sub-Saharan region exclusive of the Republic of Sudan. The Republic of Sudan was included in this study as it is the only country in Northern Africa supported by The Global Alliance for Vaccines and Immunizations (GAVI). Although South Sudan is GAVI-supported, there are no published statistics on its immunisation financing. The Sub-Saharan African nations hold about 25% of the United Nation General Assembly (7). These countries were included in this study as they had complete data available for the entire period 2013-2017. Data was retrieved from WHO Library, GAVI and World bank databases. Data for vaccine coverage were obtained for Bacille Calmette-Guerin (BCG), PCV3, Measles Containing Vaccine (MCV) and Diphtheria, Pertussis, Tetanus 3 (DPT3) from the WHO-UNICEF estimates of vaccine coverage for all the countries for the entire period. Data for fertility rate, under five mortality rates, under five population and the total population were obtained from the World Bank (7). We focused on DPT3 in this study in accordance with previous literature that showed DPT3 as the strength marker for immunisation coverage in the community (8). Immunisation financing sources in Sub-Saharan Africa include the Gross Domestic Product and GAVI.

### Statistical analysis

We performed both descriptive statistical analyses. Binary data were summarised using percentages and continuous variables using medians (IQR) given the presence of huge variations across countries. We fitted a generalised linear model to model the effect of immunisation expenditure and health expenditure (as a % of the GDP) on immunisation coverage for the different countries. Our model included the time averages of the time-varying covariates and allowed for clustering of the standard errors. The outcome variable for the model was vaccine coverage; each vaccine coverage (MCV1, DPT3 and BCG) was modelled separately. The independent variables included immunisation expenditure and health expenditure as a percentage of Gross Domestic Product (GDP). Additional variables included fertility rate, under five mortality rates, under five population and the total population. The choice of independent variables added on the model were based on the fact that they were likely to be associated with immunisation coverage. All countries without complete data were excluded from our analysis. All statistical tests were performed at the 5% level of significance. Statistical analyses were performed in R software version 3.6.3 (R Core Team, Vienna, Austria) and STATA version 15 (StataCorp, College Station, TX, USA).

## Results

### Demographic characteristics

We utilised data from twenty-four countries with complete data on immunisation coverage and health expenditure on immunisation between 2013 and 2017. **Table 1** presents a summary of the demographic characteristics of the twenty-four countries in this study. The median DPT3, BCG and PCV3 coverage were satisfactory overall, exceeding 80% over the four-year period. Of the four vaccines, immunisation coverage increased most for PCV3 and MCV1 (6.5% and 7% respectively) between 2013 and 2017. This is unsurprising as several countries first introduced the vaccine in their immunisation programme within the study period (2013-2017). The highest health expenditure (as a % of the GDP) was in 2014 (28%) and lowest in 2016 (25%). Health budget on immunisation with donor support remained constant at 0.1% across 2013 to 2017 and between 0.6 and 0.5 without donor support across 2013 to 2017. The median fertility rates were highest in 2013 (4.8) and lowest in 2016 (4.5). There was an increase in the under-five population from 2013-2016 and then a decrease in 2017 (2.07 million). Overall, there was evidence of decline in mortality rates and fertility rates between 2013 and 2017, while the population of under five years increased in the same period.

**Table 1:**
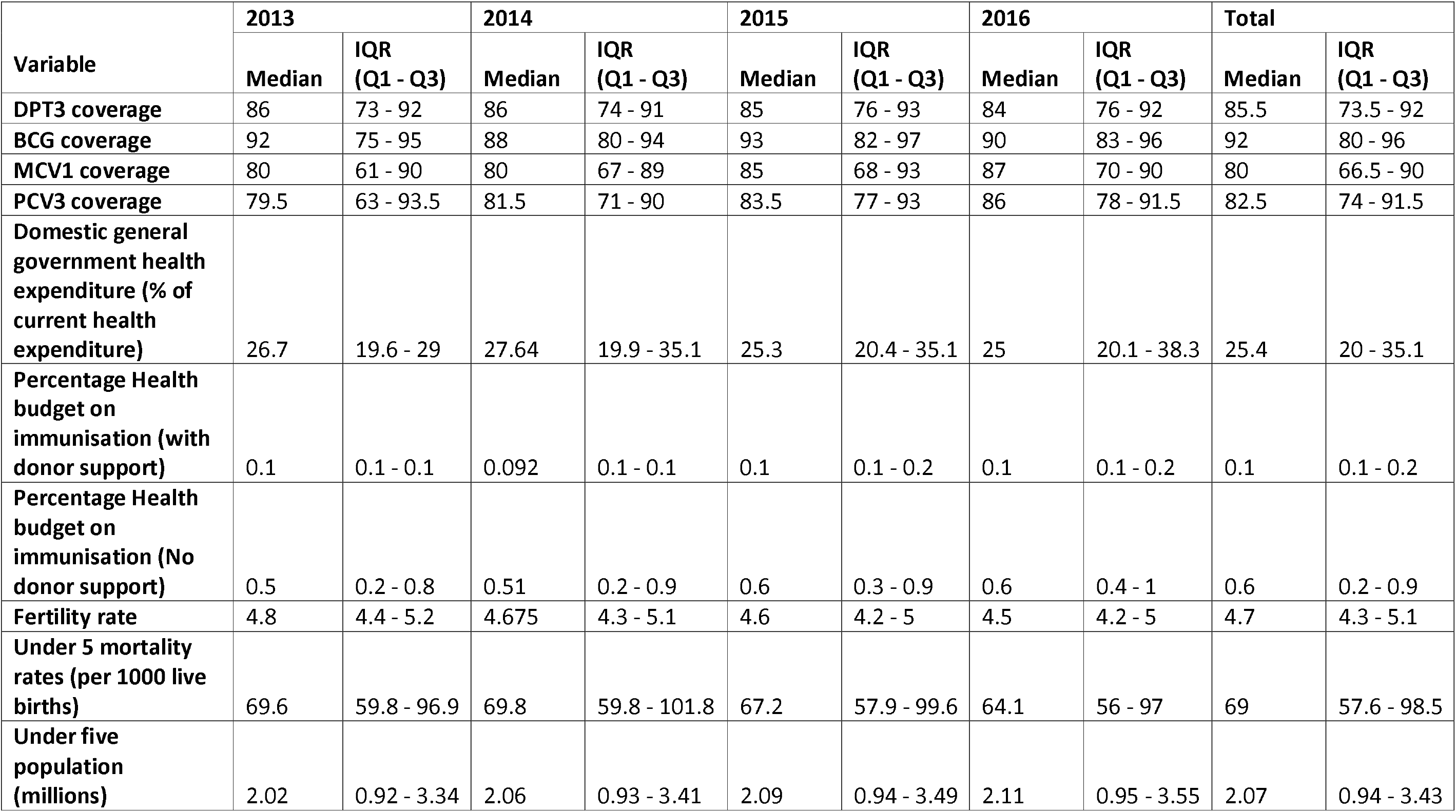
Summary of demographic characteristics of the 25 sub-Saharan countries (2013-2016)

| Variable | 2013 |  | 2014 |  | 2015 |  | 2016 |  | Total |  |
| --- | --- | --- | --- | --- | --- | --- | --- | --- | --- | --- |
|  | Median | IQR (Q1 - Q3) | Median | IQR (Q1 - Q3) | Median | IQR (Q1 - Q3) | Median | IQR (Q1 - Q3) | Median | IQR (Q1 - Q3) |
| **DPT3 coverage** | 86 | 73 - 92 | 86 | 74 - 91 | 85 | 76 - 93 | 84 | 76 - 92 | 85.5 | 73.5 - 92 |
| **BCG coverage** | 92 | 75 - 95 | 88 | 80 - 94 | 93 | 82 - 97 | 90 | 83 - 96 | 92 | 80 - 96 |
| **MCV1 coverage** | 80 | 61 - 90 | 80 | 67 - 89 | 85 | 68 - 93 | 87 | 70 - 90 | 80 | 66.5 - 90 |
| **PCV3 coverage** | 79.5 | 63 - 93.5 | 81.5 | 71 - 90 | 83.5 | 77 - 93 | 86 | 78 - 91.5 | 82.5 | 74 - 91.5 |
| **Domestic general government health expenditure (% of current health expenditure)** | 26.7 | 19.6 - 29 | 27.64 | 19.9 - 35.1 | 25.3 | 20.4 - 35.1 | 25 | 20.1 - 38.3 | 25.4 | 20 - 35.1 |
| **Percentage Health budget on immunisation (with donor support)** | 0.1 | 0.1 - 0.1 | 0.092 | 0.1 - 0.1 | 0.1 | 0.1 - 0.2 | 0.1 | 0.1 - 0.2 | 0.1 | 0.1 - 0.2 |
| **Percentage Health budget on immunisation (No donor support)** | 0.5 | 0.2 - 0.8 | 0.51 | 0.2 - 0.9 | 0.6 | 0.3 - 0.9 | 0.6 | 0.4 - 1 | 0.6 | 0.2 - 0.9 |
| **Fertility rate** | 4.8 | 4.4 - 5.2 | 4.675 | 4.3 - 5.1 | 4.6 | 4.2 - 5 | 4.5 | 4.2 - 5 | 4.7 | 4.3 - 5.1 |
| **Under 5 mortality rates (per 1000 live births)** | 69.6 | 59.8 - 96.9 | 69.8 | 59.8 - 101.8 | 67.2 | 57.9 - 99.6 | 64.1 | 56 - 97 | 69 | 57.6 - 98.5 |
| **Under five population (millions)** | 2.02 | 0.92 - 3.34 | 2.06 | 0.93 - 3.41 | 2.09 | 0.94 - 3.49 | 2.11 | 0.95 - 3.55 | 2.07 | 0.94 - 3.43 |

### Health expenditure and immunisation expenditure

**Figure 1 (Left panel)** shows the health expenditure (as a % of the GDP) and the health expenditure on immunisation (as a percentage of the total health expenditure) for 24 African countries. The World Bank 2019 data on the health expenditure (as a % of the GDP) show that there is variation in this expenditure. Liberia showed a sharp increase in the health expenditure as a percentage of GDP. Other countries with a sharp increase are Sierra Leone, Madagascar and Cote d’Ivoire. Djibouti had a sharp decline in health expenditure over the 5 years although it had the highest health expenditure with 61% in 2017 compared to Liberia which had the lowest Health expenditure at 4%. Rwanda generally faced a flat trend.

**Figure 1:** (Left panel) Health expenditure on immunisation as a percentage of the GDP; (Right panel) Health expenditure on immunisation as a percentage of the total health expenditure for 24 sub-Saharan Africa countries (2013-2017).

### Immunisation financing

**Figure 1 (Right panel)** reports the expenditure on vaccines for routine immunisation (as a proportion of total health expenditure) with and without donor support. The health expenditure on immunisation (as a percentage of total health expenditure) for all the countries showed a constant trend of below 1%. Evidently, donor support in financing the cost of vaccines accounted for a larger proportion relative to government contribution. There was limited variability in health expenditure on immunisation with the majority evidenced by the marginal increase and declines over the five-year period, except for a few countries like Central Africa Republic, Sierra Leone, and Liberia. Burkina Faso (0.14%), Burundi (0.14%), Côte D’Ivoire (0.07%), Djibouti (0.15%), Eritrea (0.11%), Ethiopia (0.27%), Gambia (0.19%), Ghana (0.07%), Madagascar (0.04%), Mozambique (0.22%), Rwanda (0.06%), Sierra Leone (0.12%), Sudan (0.08%), Zambia (0.07%), Zimbabwe (0.14%), Togo (0.65%), Senegal (0.04%), Guinea (0.09%), Liberia(0.15%), CAR (0.07%), Chad (0.11%),Republic of the Congo (0.01%) and Benin (0.06%). For all these countries, co-finance routine vaccines are supported by GAVI. Co-finance means that apart from GAVI financing the vaccines, the countries also contribute to the cost of GAVI-supported vaccines (9).

Additionally, **Figure 1 (Right panel)** reports how much countries would need to finance vaccines without GAVI’s support. Sierra Leone would need to contribute 1.54% in 2013, 1.90% in 2014, 2.02% in 2015, 2.32% in 2016 and 1.68% more in 2017. Similarly, the Central African Republic which had the biggest difference would need 0.84% in 2013, 3.24% in 2014, 4.48% in 2015, 2.59% in 2016 and 1.28% in 2017 more to self-finance vaccines. On the other hand, Republic of the Congo, Ghana, Djibouti, and Benin showed they were on pace to self-financing vaccines.

### Coverage trends

**Figure 2** shows the coverage of BCG, MCV1, DPT3 and PCV3 vaccines in 24 countries from 2013-2017. Zambia and Zimbabwe had missing data on BCG coverage while Guinea and Liberia, Republic of the Congo, Eritrea, Central Africa Republic, Chad had missing data on PCV3 Coverage.

**Figure 2:** Vaccine coverage for BCG, DPT, MCV1 and PCV3 between 2013 and 2017 for twenty-four sub-Saharan African countries

Most countries had relatively consistent immunisation performance as measured by the DPT3 coverage. Countries that achieved the GAVI goal of 90% coverage by 2015 included Burkina Faso (91%), Gambia (97%), Ghana (98%), Burundi (93%), Rwanda (98%), Zambia (90%), Eritrea (95%) and Sudan (93%) while Côte D’Ivoire, Mozambique, Senegal, Sierra Leone, Togo, and Zimbabwe had above 80% DPT3 coverage between 2013 to 2017. In contrast, Benin, Liberia, Madagascar, Mali, Central African Republic, Republic of the Congo, Chad, Djibouti, and Guinea were below the Global Vaccine Action Plan (GVAP) target percentage of greater than 80% DTP3 coverage. In Benin, Liberia, Madagascar and Mali the coverage was above 75%. Guinea did not acquire a 50% DPT3 coverage by the end of 2017 while coverage for MCV1 also lagged. Republic of the Congo saw huge drops between 2013 and 2016 and an increase in 2017. The Central African Republic and Guinea have struggled to bring its coverage above 50% throughout the 5 years. Chad has shown low coverage with a highest of 42% and lowest of 39%. Chad and the Central African Republic had the lowest DPT3 coverage by 2017. Although Ethiopia’s coverage showed lower immunisation coverage (below 75%) there is a significant increase in the coverage over the 5 years with a lowest of 59% and highest of 69%.

**Figure 2** shows PCV coverage. Among the 24 GAVI-eligible countries, 18 of them had adopted the PCV vaccine. These include Benin, Burkina Faso, Burundi, Côte D’Ivoire, Djibouti, Ethiopia, Gambia, Ghana, Madagascar, Mali, Mozambique, Rwanda, Senegal, Sierra Leone, Sudan, Togo, Zambia and Zimbabwe. The other countries, Central Africa Republic, Chad, Republic of the Congo, Eritrea, Guinea, and Liberia had lagged behind in the introduction of the PCV by the year 2017.

### Predicting immunisation coverage

**Table 2** shows that health expenditure on immunisation (as a proportion of total health expenditure) is not a significant predictor of immunisation coverage. Contrary to expectation, the relationship is negative. Further, it is surprising that certain countries with low expenditure on immunisation had high coverage (>80%) for example; Republic of the Congo had 90% DPT3 coverage in 2014 with a health expenditure on immunisation (as a proportion of total health expenditure) of 0.03% while Sierra Leone which had 83% DPT3 coverage and a health expenditure on immunisation (as a proportion of total health expenditure) of 0.4% in the same year. On the hand, other countries with similar expenditure on immunisation in 2017 showed different immunisation coverage. For example, we observed that Chad and Eritrea had a 41% and 95% DPT3 coverage yet they had similar health expenditure on immunisation (as a proportion of total health expenditure) of 0.10% in 2017. This similar observation was made for the second model with the variable immunisation without donor support. The inverse relationship observed for certain countries is a major contributor to the negative association observed in our model. However, we observe that the association between total health expenditure as a percentage of GDP and immunisation coverage showed a stronger link with BCG (p=0.047) and DPT3 (p=0.013) vaccines. Therefore, health expenditure as a percentage of GDP is an important predictor of immunisation coverage. Under five mortality was shown to be associated with DPT3 coverage.

**Table 2:** Model results.

|  | DPT Coverage |  |  | BCG Coverage |  |  | MCV Coverage |  |  |
| --- | --- | --- | --- | --- | --- | --- | --- | --- | --- |
|  | Beta | SE | p | Beta | SE | p | Beta | SE | p |
| Health expenditure as a percentage of GDP | 0.0152 | 0.006 | 0.0130 | 0.0182 | 0.00917 | 0.0470 | 0.0092 | 0.007 | 0.210 |
| Health expenditure on immunisation | -0.3770 | 0.4136 | 0.3620 | -0.8876 | 0.65870 | 0.1780 | -0.0829 | 0.341 | 0.808 |
| Fertility rate | -2.4677 | 1.5933 | 0.1210 | -2.6960 | 2.07833 | 0.1950 | -5.63E-01 | 1.592 | 0.723 |
| Mortality rate among under 5 years old | -0.0020 | 0.0007 | 0.0080 | -0.0013 | 0.00138 | 0.3280 | -6.25E-04 | 0.001 | 0.580 |
| Under 5 population | 1.37E-06 | 0.0000 | 0.1040 | 1.87E-06 | 1.21E-06 | 0.1230 | 4.26E-07 | 6.89E-07 | 0.537 |
| Population | -1.17E-07 | 7.31E-08 | 0.1110 | -1.96E-07 | 1.12E-07 | 8.00E-02 | -5.03E-08 | 6.17E-08 | 0.415 |
| **Model with expenditure on health immunisation without donor support** |  |  |  |  |  |  |  |  |  |
| Health expenditure as a percentage of GDP | 0.0148 | 0.0059 | 0.0110 | 0.0170 | 0.0091 | 0.0610 | 0.0061 | 0.0069 | 0.380 |
| Health expenditure on immunisation | -0.0630 | 0.1325 | 0.6350 | -0.1483 | 0.2025 | 0.4640 | -0.1192 | 0.1375 | 0.386 |
| Fertility rate | -2.4965 | 1.6513 | 0.1310 | -2.7822 | 2.1672 | 0.1990 | -0.7128 | 1.6186 | 0.660 |
| Mortality rate among under 5 years old | -2.05E-03 | 7.88E-04 | 0.0090 | -0.0014 | 0.0013 | 0.2660 | -0.0004 | 0.0010 | 0.674 |
| Under 5 population | 1.17E-06 | 9.14E-07 | 0.1990 | 1.39E-06 | 1.34E-06 | 0.2980 | 3.82E-07 | 7.30E-07 | 0.601 |
| Population | -1.04E-07 | 7.36E-08 | 0.1600 | -1.61E-07 | 1.18E-07 | 1.73E-01 | -5.53E-08 | 6.07E-08 | 0.363 |

### Predicting immunisation coverage

We sampled countries with the lowest percentage of DPT3 and BCG coverage to predict immunisation coverage as we vary total health expenditure (as a % of the GDP). The results are visualised in **Figure 3** and **Figure 4. Figure 3** shows immunisation coverage for countries with less than desired level of DPT3 coverage (below 80%) to visualise how much expenditure on health as a percentage of GDP would result in an increase in DPT3 coverage with all other variables unchanged. These countries include Benin, Central African Republic, Chad, and Guinea. We used the health expenditure (as a % of the GDP) variable to perform the prediction as it was the most important variable of interest associated with immunisation coverage.

**Figure 3:** Predicted (DPT3) immunisation coverage whilst varying health expenditure on immunisation as a percentage of GDP

**Figure 4:** Predicted (BCG) immunisation coverage whilst varying health expenditure on immunisation as a percentage of GDP

On average, we observe a steady increase in the predicted immunisation coverage for the four countries as health expenditure (as a % of the GDP) increases. For Benin and Guinea, to realise a desired level of DPT3 coverage of at least 80%, we notice that the 2017 total health expenditure should be increased to about 45% of the total GDP. In Chad and the Central Africa Republic, a 45% increase in health expenditure (as a percentage of GDP) would result in a 60% coverage in DPT3 coverage. Notably, these predictions assume other predictor variables in the original model remain unchanged. Although that assumption may not hold, our aim was to show how health expenditure would realise a desired level of coverage given the known values of other important variables.

Countries that achieved a 90% DPT3 coverage such as Burkina Faso, the Gambia and Sudan spent about 43%, 23%, and 46% respectively on health expenditure (as a percentage of GDP). For countries that achieved a 80% DPT3 coverage such as Liberia, Mozambique and Togo the health expenditure as a percentage of GDP was 17%, 21%% and 15% respectively

**Figure 4** shows immunisation coverage for countries with less than desired level of BCG coverage (below 80%). These countries include Ethiopia, Central African Republic, Mali, and Guinea. We predict that increasing the 2017 health expenditure (as a percentage of GDP) by 25% to 35% would result in 80% coverage in BCG in these countries.

## Discussion

In this study, we examined the influence that immunisation expenditure and health expenditure (as a % of the GDP) has on immunisation coverage between 2013 and 2017. Our study findings showed that over the five-year period, immunisation coverage for DPT3, BCG and PCV3 was satisfactory overall, with median values exceeding 80%. This reflects progress towards increasing access to immunisation and a potential for a decrease in under-five mortality from vaccine-preventable diseases. While the recorded high immunisation coverage for DPT3, BCG and PCV3 are noteworthy, the GAVP target of 90% coverage was not attained with many countries recording suboptimal national and sub-national immunisation coverage amid struggle to increase immunisation financing and vaccine accessibility. From our study result, between 2013 and 2017, immunisation coverage increased mostly for PCV3 and MCV1 by 6.5% and 7% respectively. This increased coverage may be attributed to the early operational and programmatic readiness at national and subnational levels, with high-level political interest at the global and national level (10). However, it was suggested that there could have been rate-limiting factors to the high-achieved PCV3 coverage which are not limited to low capacity of health care workers, poor vaccine management, and poor coordination at management level (10). A recent study identified weak health systems, lack of commitment, inadequate quality social mobilisation and community engagements, and planning capacity among others as factors that limit vaccination coverage. Additionally, the introduction of new vaccines in developing contexts may be facilitated through innovations in the supply chain and new digital technologies for training, communication, and research.

While 14 countries achieved the GAVI goal of 90% DPT3 coverage by 2015, countries like Benin, Central African Republic, Chad, and Guinea still had the lowest (below 50%) DPT3 immunisation coverage by 2017. Similar results have been reported by Mosser et al (11) and Ikilezi et al (12) where low DPT3 coverage was observed in Central African Republic, Chad and Guinea in 2016. In Guinea, the low coverage has been attributed to low maternal education attainment (13), other possible explanations could be the low investment in health expenditure (as a % of the GDP) in these countries. This cannot be generalised as the main contributing factor for the low coverage as other countries like Sierra Leone had one of the lowest allocations of health expenditure (as a % of the GDP), yet they attained higher DPT3 coverage. Therefore, other factors such as conflict (14), vaccine shortages (15), and inadequate knowledge on vaccination by health workers (15) could be attributed to the observed low immunisation in these countries during this period.

Our study results highlighted that 21 of the 24 countries by 2017 financed less than half of their vaccines programme with the remainder financed by GAVI. These findings are relatively lower compared to Lydon et al (16) findings where government spending on vaccines in Africa were estimated to range between 48% and 53% between 2000 and 2006. Another study reported similar findings to ours where 28 countries in Africa were financing less than half of their immunisation budget by 2017 of which 10 were funding less than 20% (17). According to the study only 5 countries (not included in our analysis) financed 100% of their immunisation budget (17). Most countries in Sub-Saharan Africa are still and perhaps more heavily reliant on GAVI and other external support to finance more than half of their vaccination programmes. It is imperative that countries in Sub-Saharan Africa should explore innovative health financing options and contextually appropriate domestic resource mobilisation strategies that will foster a significant and sustained increase in the domestic funding for immunisation programmes. Increased domestic funding of immunisation programmes in Sub-Saharan Africa is important to prevent the funding gap that may result as development assistance for health dwindles and some countries transition away from donor funding.

Our findings showed health expenditure on immunisation (as a proportion of total health expenditure) is not a significant predictor of immunisation coverage. Contrary to expectation, we observed a negative relationship. Certain countries with low expenditure on immunisation had high coverage (>80%). On the other hand, countries with similar health expenditure (as a % of the GDP) had a different impact on immunisation coverage for DPT3 vaccine. For instance, in 2017, Chad and Eritrea both had similar health expenditure (as a % of the GDP) of 24% while their immunisation coverage was 41% and 95% DPT3 respectively. The observed difference in immunisation coverage despite a similar level of expenditure could be linked to other factors that are peculiar to each country that could affect projected immunisation coverage. For example, it is likely that the internal security challenges in Chad (14) could have affected the systems for cold chain, vaccine storage and delivery and scared mothers from taking their children for immunisation thus resulting in the recorded immunisation coverage of 41%. On the other hand, certain countries with low expenditure on immunisation had high coverage (>80%) for example, Republic of the Congo and Sierra Leone. This low expenditure on immunisation could have been influenced by the low fiscal space for health, low prioritisation of immunisation by the government and a possible substitution effect - with government reallocating resources for immunisation to other priorities because of the availability of external funding for healthcare from organisations like GAVI and UNICEF. The low fiscal space for health is linked to the macroeconomic realities in these countries as countries would likely spend more on health if they had higher GDP, provided health remained a key priority for the government. Also, it is important to note that while significant amounts of immunisation spendings are on supplementary immunisation activities (SIAs campaign) etc, the outcome of these exercises most times are not added to routine immunisation coverages. This may have also accounted for the reason why our analysis did not find any positive correlation between health expenditure on immunisation (as a % of total health expenditure) and immunisation coverage that suggest that the former could be a strong predictor of the latter.

The analysis result further showed an association between DPT3 coverage and under-5 years mortality. As expected, within the 5-year period explored by our study, under-5 mortality decreased as DPT3 coverage increased. Although our study did not find any statistically significant relationship between health expenditure as a percentage of GDP and immunization coverage, there is a possibility that increases in the former maybe linked to the observed decline in under-5 mortality. Cardona M et al (2022) postulated an association between child mortality and GDP, showing that a 5% reduction in GDP per capita in 2020 was estimated to cause an additional 282,996 deaths in children under 5 years in 2020. A study conducted by Onishchenko et al (2019) suggested that increases in national immunisation expenditure correlated with reduced infant mortality and increased life expectancy. It is to be noted that the selected countries in Onishchenko et al study are not among the United Nations Development Program’s sub-Saharan African countries; the two North African countries (Egypt and Morocco) that were included are 100% financed by their respective governments. It is globally presumed that decline in under-5 years mortality trend is one of the major expected outcomes of health spending on immunisation. We hypothesise that expenditure on some sub-sectors of national health programmes (such as vertical community-based management of acute malnutrition, integrated management of neonatal and childhood illness (IMNCI), integrated community case management (iCCM) etc.) implemented in integration with immunisation could have contributed to the reduced mortality rate observed. In that context, the positive impact of health expenditure as a percentage of GDP on under-5 years mortality in our study should not be interpreted as an inevitable outcome regardless of how the money is spent as studies have suggested that the quality of national institutions (degree of public sector accountability, stability of the political system, workforce productivity and so forth) can influence the effectiveness of public spending.

The health expenditure on immunisation in most of the countries was below 1% between 2013-2017, with variations observed among countries. For example, in Mali, Togo, Zimbabwe and Sierra Leone, we observed that the health expenditure on immunisation (as a percentage of the total health expenditure) was above one percent. The health expenditure on immunisation of 1% was lower than the 2% average seen in other LMIC economies outside Africa. This higher expenditure on immunisation may indicate that immunisation was prioritised in these countries, with increased funding from domestic and external sources. Immunisation is primarily financed through government expenditure such as government revenue (tax), borrowing and grants. Government revenues increase as the economy of the country grows. Therefore, economic downtowns lead to low expenditure requiring governments to prioritize budgets allocations between competing sectors. The Sabin program rolled out in Mali and Sierra Leone, may have influenced immunization coverage as it focused on linking the agenda of the ministry of health, ministry of finance and the parliament, supporting collective actions and resulting in a government owned sustainable immunisation program. In addition, the program helped these countries compare plans for developing national immunisation trust funds such as decentralisation, legislation and budget reforms resulting in improved budget allocation (18).

The highest health expenditure (as a % of the GDP) was in 2014 (28%) and lowest in 2016 (25%). Health budget on immunisation with donor support remained constant at 0.1% across 2013 to 2017 and between 0.6 and 0.5 without donor support across 2013 to 2017. It is likely that the predominance of donor funding for immunisation in most of the countries may account for the near constant level of funding on immunisation as support for immunisation from donors is more predictable unlike domestic funding from Government which may be subject to change from one budget cycle to another. Health expenditure (as a % of the GDP) could have been reduced because of a shift in government’s priorities with resources that should have otherwise been spent on the health sector being allocated to other sectors. Furthermore, our prediction model showed that more than 45% increase in health expenditure as a percentage of GDP would be needed to meet the GAVI target of 80% in the countries with the least DPT3 coverage. This implies that there is dire need for these governments to review their health policies and adopt legislations that target equitable protection of the health of all its citizens, such as in the case of Burkina Faso (19). The involvement of the communities and civil societies in immunisation programmes could also drive success in these countries (17).

Our study finding that a steady increase in the predicted immunisation coverage as health expenditure (as a % of the GDP) increased implies that for countries to realise a desired level of DPT3 coverage, a minimum level of increase in health investment/expenditure would be required. These predictions assume other predictor variables in the original model remain unchanged. Although it is unlikely that all other predictor variables can be kept constant,, our model was designed to show how health expenditure as a single variable would affect immunization coverage if other important variables were known and kept constant. Countries are advised to sustain and increase their spending on health. A study conducted by Moreno-Serra and Smith (2011) on the Effects of Health Coverage on Population Outcomes offers hard evidence that investing in broader health coverage can generate significant gains in terms of population health. Furthermore, the effective implementation of integrated primary health care has also been reported to have contributed to improved health coverage including immunisation coverage. Impact of spending on other components of the PHC such as nutrition could produce an indirect positive effect on immunisation outcome.

There are some limitations to this study. First, while our secondary data was sourced from reliable institutions including GAVI, WHO and other UN agencies, certain estimates such as those of vaccine coverage are subject to sampling error and information bias, and other differences in data collection methods in surveys across countries. Secondly there is limited availability of further detailed information on vaccine coverage, such as the distribution of vaccines across socio-economic groups which limits further exploration on whether the way immunisation is finance may affect coverage across various population subgroups. Despite this, we believe our findings provide an important starting point when discussing immunisation financing in low- and middle-income countries. An important caveat is that our results do not represent causal relationships between the given predictors and immunization coverage.

### Recommendation for action plans/strategies for Africa

We recommend that countries with the least DPT3 and BCG coverage rates should increase their health expenditure as a percentage of GDP by 45% and 35% respectively to achieve at least DPT 3 and BCG coverage of 80%. This steady increase will move their expenditure on health as a % of GDP closer to global LMICs average of about 2% of GDP. Since DPT3 coverage is found to be associated with under5 mortality and expenditure on health as a % of GDP, such increased spending on health could also translate into reduction in under5 mortality in those countries. In the allocation of resources, prioritising expenditure on health as % of GDP should be a concern of the government due to its potential impacts on population health. Also, sustainability of financing is important to achieve impactful health outcomes. While sustained support from donors remains vital, governments across these countries should prioritise spending on health as a % of GDP in their fiscal planning and implementations, including resource mobilisation. Donor support remained at a relatively constant ratio during the period the study covered. Government and non-governmental bodies needed to ramp up resources to meet the immunisation gaps, by exploring evidence-based strategies and best practices in financing national health programmes. Considering the positive externality associated with immunisation, promoting and/or strengthening implementation of *integrated health care delivery*, could enhance efficiency and effectiveness. Also, countries may from time to time conduct cost-effectiveness analysis of their intervention strategies related to health, especially for immunisation.

Globally, the concept of value for money has continued to gain more acceptance in the management of allocated humanitarian and development funds. This study finding did not support the assumption that health expenditure on immunisation is a predictor of immunisation coverage, and it may be beneficial for government and non-government stakeholders to strengthen further accountability systems around immunisation programmes including adoption of value for money systems. Countries should continue to explore immunisation investment alternatives, and balance competing objectives in order to derive maximum benefits. Also, countries should prioritise accountability and keep records of outcomes of immunisation interventions such as SIAs whose data are often not incorporated into routine immunisation coverage and annual performance reporting in many countries. Furthermore, we recommend that other metrics should be in place in addition to coverage to track the success of health expenditure on immunisation, considering the externalities that could affect outcomes. This aligns with what is proposed in Immunisation agenda 2030 relating to immunisation coverage target and data management.

Also, in view of the consistent mention of the negative impact of conflicts on immunisation and health outcomes in some countries, we recommend that countries in conflicts adopt strategies that would guarantee health access to all parties in conflicts as this will optimize vaccine access and/or availability. Constant engagement by state and non-state actors is critical to having access, and health impacts. Immunisation interventions and spendings should be tailored in the direction that accommodate local context, with relative guarantee for good outcomes. Allocation of resources should be justified and effectively deployed, and it should target significantly the neglected and vulnerable population to ensure no one is left behind. Finally, we recommend that countries should continue to generate and keep detailed immunisation data at national and subnational levels including data on investments, outcomes, and challenges to guide and improve intervention by government and partners in the future. Documenting systematically and in detail the economic, health and child development data around immunisation could help policy and health managers take informed planning and intervention decisions to improve outcomes.

## Data Availability

All data produced in the present study are available upon reasonable request to the authors.

## Authors’ contributions

All authors equally contributed to the writing of the manuscript. IOI conceptualised this study, conducted the data collection and management, planned the methodology, and contributed to the data analysis. JT participated in the conceptualisation of the study, contributed to data collection and management, and contributed to the data analysis. LO participated in the conceptualisation of the study, participated in the data management, planned the methodology, and solely conducted the data analysis. All authors read and approved the final manuscript.

## Conflict of interest

The authors declare no conflict of interest.

## Acknowledgements

We thank Dkhimi Fahdi for his useful comments to improve this manuscript.

## Funding statement

This study did not receive any funding

## Data availability statement

All data produced in the present study are available upon reasonable request to the authors.

## Footnotes

1 Domestic general government health expenditure (% of current health expenditure) indicates how much resources is the public sector devoting for health. Public sources include domestic revenue as internal transfers and grants, transfers, subsidies to voluntary health insurance beneficiaries, NPISH or enterprise financing schemes as well as compulsory prepayment and social health insurance contributions.

## Notes

### Competing Interest Statement

The authors have declared no competing interest.

## References

1. Ministerial Conference on Immunization in Africa. Addis Declaration on Immunization — Ministerial Conference on Immunization in Africa [Internet]. [cited 2021 Jun 5]. Available from: http://immunizationinafrica2016.org/ministerial-declaration-english

2. Immunization | WHO | Regional Office for Africa [Internet]. [cited 2021 Apr 29]. Available from: https://www.afro.who.int/health-topics/immunization

3. Maternal and Neonatal Tetanus | WHO | Regional Office for Africa.

4. De Roeck D, AbtAssociates Inc. Review of Financing of Immunization Programs in Developing and Transitional Countries. Bethesda, MD: Partnerships for Health Reform Project, Abt Associates Inc.; Dec p. xiii. Report No.: 12. 1998;

5. WHO. Ensuring Sustainability: Immunization Financing in an Era of Transition [Internet]. 2016 [cited 2021 Jun 5]. Available from: https://www.afro.who.int/sites/default/files/2017-12/MCIA Brief_Immunization Financing in an Era of Transition.pdf

6. World Bank Country and Lending Groups [Internet]. [cited 2022 Apr 7]. Available from: https://datahelpdesk.worldbank.org/knowledgebase/articles/906519-world-bank-country-and-lending-groups

7. W101. Sub-Saharan Africa | Geopolitics [Internet]. 2021 [cited 2021 Jun 5]. Available from: https://world101.cfr.org/rotw/africa/geopolitics.

8. Arevshatian L, Clements CJ, Lwanga SK, Misore AO, Ndumbe P, Seward JF, Taylor P. An evaluation of infant immunization in Africa: Is a transformation in progress? Bull World Health Organ. 2007;85.

9. GAVI. Co-financing policy [Internet]. [cited 2022 Apr 7]. Available from: https://www.gavi.org/programmes-impact/programmatic-policies/co-financing-policy

10. Olayinka F, Ewald L, Steinglass R. Beyond new vaccine introduction: the uptake of pneumococcal conjugate vaccine in the African Region. Pan Afr Med J. 2017;27.

11. Mosser JF, Gagne-Maynard W, Rao PC, Osgood-Zimmerman A, Fullman N, Graetz N, Burstein R, Updike RL, Liu PY, Ray SE, et al. Mapping diphtheria-pertussis-tetanus vaccine coverage in Africa, 2000–2016: a spatial and temporal modelling study. Lancet. 2019;393.

12. Ikilezi G, Augusto OJ, Sbarra A, Sherr K, Dieleman JL, Lim SS. Determinants of geographical inequalities for DTP3 vaccine coverage in sub-Saharan Africa. Vaccine. 2020;38.

13. Bado AR, Susuman AS. Women’s education and health inequalities in under-five mortality in selected sub-saharan African countries, 1990-2015. PLoS One. 2016;11.

14. Grundy J, Biggs BA. The impact of conflict on immunisation coverage in 16 countries. Int J Heal Policy Manag. 2019;8.

15. Ogbuanu IU, Li AJ, Anya B philomene M, Tamadji M, Chirwa G, Chiwaya KW, Djalal MEH, Cheikh D, Machekanyanga Z, Okeibunor J, et al. Can vaccination coverage be improved by reducing missed opportunities for vaccination? Findings from assessments in Chad and Malawi using the new WHO methodology. PLoS One. 2019;14.

16. Lydon P, Beyai PL, Chaudhri I, Cakmak N, Satoulou A, Dumolard L. Government financing for health and specific national budget lines: The case of vaccines and immunization. Vaccine. 2008.

17. Mihigo R, Okeibunor J, Anya B, Mkanda P, Zawaira F. Challenges of immunization in the African Region. Pan Afr Med J. 2017;27.

18. McQuestion M, Gnawali D, Kamara C, Kizza D, Mambu-Ma-Disu H, Mbwangue J, de Quadros C. Creating sustainable financing and support for immunization programs in fifteen developing countries. Health Aff. 2011;30.

19. Universal Health Coverage Partnership. A historic step towards Health for All: Burkina Faso’s new universal health insurance law | Universal Health Coverage Partnership [Internet]. 2018 [cited 2022 Apr 7]. Available from: https://www.uhcpartnership.net/an-historic-step-towards-health-for-all-burkina-fasos-new-universal-health-insurance-law-3/

